# Long-term and Landmark Analysis of Transcatheter versus Surgical Aortic-Valve Replacement in Severe Aortic Stenosis

**DOI:** 10.1101/2023.12.22.23300476

**Authors:** Xiaowen Zhang, Lina Kang, Lian Wang, Kun Wang, Wei Xu, Biao Xu, Xinlin Zhang

## Abstract

**Background:** Previous reports of long-term outcomes of TAVR focus on higher risk patients and suggest potential temporal changes. The indications of TAVR have expanded to low-risk patients. We aimed to evaluate the long-term and temporal performances of transcatheter aortic valve replacement (TAVR) compared to surgical aortic valve replacement (SAVR).

**Methods:** Randomized controlled trials reporting outcomes with at least 1-year follow-up were included. The primary outcome was the composite of all-cause death or disabling stroke.

**Results:** We included 8 trials with 8,749 patients. TAVR was associated with a higher risk of long-term (5-year) primary outcome compared to SAVR among higher-risk (odds ratio [OR], 1.25; 95% CI, 1.07–1.47) but not lower-risk participants (1.0 [0.77–1.29]). However, a significant temporal interaction was detected in both risk profiles. TAVR with balloon-expandable valves was associated with a higher risk of long-term primary outcome compared to SAVR (1.38 [1.2–1.6]), whereas no statistical difference was found with self-expanding valves (1.03 [0.89–1.19]). There was a significant interaction between the two valve systems, and a temporal interaction was detected in both systems. Overall landmark analysis revealed a lower risk in TAVR within the initial 30 days (0.76 [0.6, 0.96]), comparable between 30 days to 2 years (1.04 [0.85, 1.28]), and higher beyond 2 years (1.36 [1.15– 1.61]). Analysis for all-cause death generated largely similar results.

**Conclusions:** TAVR was associated with a higher long-term risk of primary outcome compared to SAVR in higher-risk patients and with balloon-expandable valves. However, a characteristic temporal interaction was documented in all subgroups. Future studies are warranted to test these findings.

## Background

Transcatheter aortic valve replacement (TAVR) has emerged as a popular treatment for patients with severe aortic stenosis, surpassing surgical procedures in some countries [1]. We previously indicated a potential higher mortality associated with TAVR compared to surgical aortic valve replacement (SAVR) at 5-year follow-up [2], mainly in high risk patients [3–5]. The long-term performance of TAVR versus SAVR in patients with lower risk remains uncertain. Additionally, the temporal changes in TAVR performance at different timepoints have yet to be determined. Given the expansion of TAVR to low-risk patients with increased life expectancy, this assessment holds critical clinical importance.

The 5-year follow-up data from nearly all registered comparative randomized controlled trials (RCTs) of TAVR vs SAVR have recently been published [6–9]. We therefore are able to assess the long-term outcomes of TAVR and conduct a landmark analysis to identify the timepoint at which the performance of TAVR might diverge from SAVR, as indicated in some studies [5]. We undertook an updated meta-analysis of RCTs to evaluate the long-term and temporal performances of TAVR compared to SAVR, both overall and within important subgroups.

## Methods

We reported the meta-analysis in accordance with the Preferred Reporting Items for Systematic Reviews and Meta-Analyses (PRISMA) guideline (Table S1).

### Data Sources and Searches

PubMed, the Cochrane Central Register of Controlled Trials, EMBASE, and major conference proceedings were systematically searched from inception through October 25, 2023, an update of our previous meta-analysis [2]. The computer-based searches combined terms and keywords which included transcatheter aortic valve implantation, transcatheter aortic valve replacement, TAVI, TAVR, and randomized trial. Two investigators independently hand-searched the references of identified studies and relevant reviews to identify any additional relevant trials.

### Study Selection

Two reviewers (X.W.Z. and X.L.Z.) conducted independent screening of titles and abstracts to determine eligibility of the studies. Full-text articles were retrieved for studies that were deemed potentially relevant. In cases where discrepancies arose, a third investigator (L.K.) resolved the discrepancies. Eligible studies had to be RCTs evaluating TAVR vs SAVR in patients with severe aortic stenosis, and reporting outcomes of interest with at least 1-year follow-up. Nonrandomized observational studies, studies comparing different types of TAVR devices, and studies with less than 1-year follow-up were excluded.

### Outcome Measures

The primary outcome was the composite of all-cause death and disabling stroke. Secondary outcomes included all-cause death, cardiovascular death, myocardial infarction, stroke, transient ischemic attack (TIA), major bleeding, major vascular complications (MVC), permanent pacemaker implantation (PPM), new-onset atrial fibrillation, aortic-valve reintervention, rehospitalization, and moderate or severe paravalvular leak (PVL).

### Data Extraction and Quality Assessment

Two investigators (X.W.Z. and X.L.Z.) independently extracted the data using a pre-specified form. Whenever possible, data from the intention-to-treat analysis were extracted; otherwise, data from the as-treated analysis were extracted. The same investigators also assessed the risk of bias in the included RCTs using the Cochrane Risk of Bias 2.0 tool.

### Statistical Analysis

Summary measures were reported as odds ratios (ORs) and pooled using random-effects models (DerSimonian–Laird method). Data were analyzed separately for different time points, including data within 30 days, 1 year, 2 years, and 5 years (one trial reported 4-year outcome and was used), and categorized as early, short-term, midterm, and long-term outcomes, respectively. Landmark analysis was also conducted for intervals within 1 year, between 1 year and 2 years, and beyond 2 years.

Events occurring within 1 year were further divided into events within 30 days and events between 30 days and 1 year to further explore the timing of performance change. For trials in which only one of the arms had no events, the 0.5 continuity correction was applied. Stratified analyses were performed based on surgical risks (higher and lower risks) and TAVR systems (balloon-expandable valves [BEV] and self-expanding valves [SEV]). The higher-risk group included trials involving patients with extreme, high, and intermediate-to-high surgical risk, while the lower-risk group included trials involving patients with low and low-to-intermediate risk, as determined by the evaluation using the Society of Thoracic Surgeons predicted risk of mortality (STS-PROM) score. Between-subgroup differences were assessed using the χ2-test for heterogeneity. Sensitivity analysis was performed for the primary outcomes using Hartung-Knapp-Sidik-Jonkman variance correction, and by removing an individual trial each time. Heterogeneity was evaluated using the Q and *I^2^* statistics. Several tests for publication bias were conducted, but no significant results were found. All meta-analyses were performed using Stata software version 16.0, and the Review Manager version 5.3. A 2-tailed p value <0.05 was considered statistically significant.

## Results

### Study Selection and Characteristics

We included 8 trials and 14 secondary reports that provided eligible data from these trials [3–23], involving a total of 8,749 patients (Figure S1). All 8 trials reported outcomes at 30 days and 1 year, 7 reported 2-year outcomes, one reported 4-year outcomes [8], and 6 reported 5-year outcomes [3–6, 9, 20]. The mean age was 79.2 years and 57.4% were male. Based on STS-PROM risk score, 4 trials were categorized as lower-risk trials, while the other 4 categorized as higher-risk trials. BEV was used in 3 trials, SEV in 4 trials, and a mixed TAVR system in one trial. Baseline characteristics are presented in Table S2 and Table S3. Blinding of participants and personnel was not feasible in any of the trials, but the overall risk of bias was considered low (Table S4).

### Primary Outcome

TAVR demonstrated a lower rate of primary outcome compared to SAVR at 30 days (odds ratio [OR], 0.76 [95% CI 0.6–0.96]) and 1 year (0.83 [0.72–0.96]). However, at long-term follow-up, TAVR was associated with a higher risk (1.17 [1.01–1.36]) (Figure 1). Landmark analysis indicated a significant benefit of TAVR within the first year, comparable events between 1 year and 2 years (1.19 [0.95– 1.49]), but a significant disadvantage beyond 2 years (1.36 [1.15–1.61]), with a significant temporal interaction (p for interaction<0.0001) (Figure 2). The most notable benefit of TAVR was observed within the initial 30 days, whereas no significant difference was found between 30 days and 1 year (0.9 [0.74–1.08]) (Table S5).

**Figure 1.**
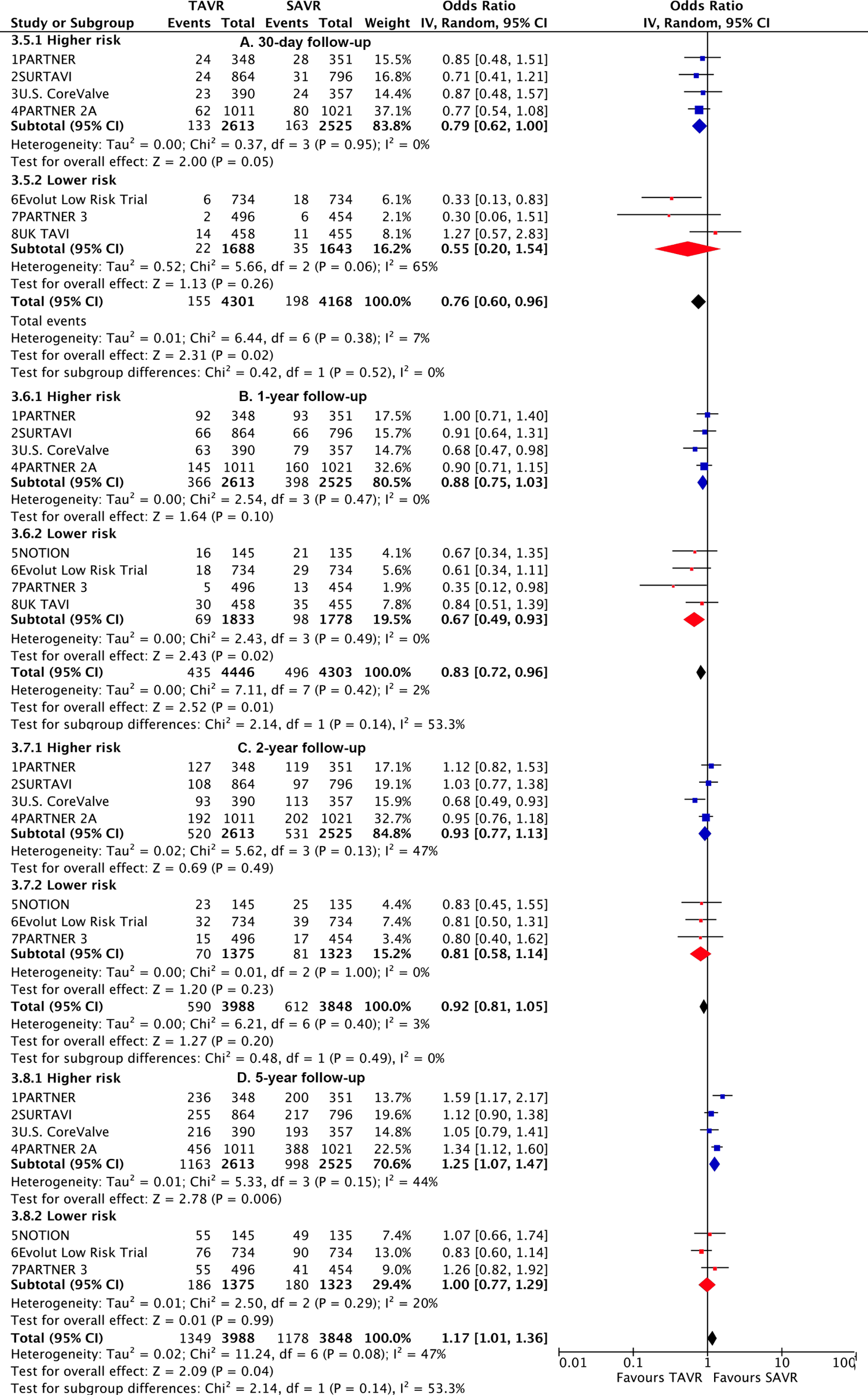
Risk estimates of all-cause death or disabling stroke for TAVR vs SAVR stratified by surgical risks at different lengths of follow-up. TAVR: transcatheter aortic valve replacement; SAVR: surgical aortic valve replacement (SAVR).

**Figure 2.**
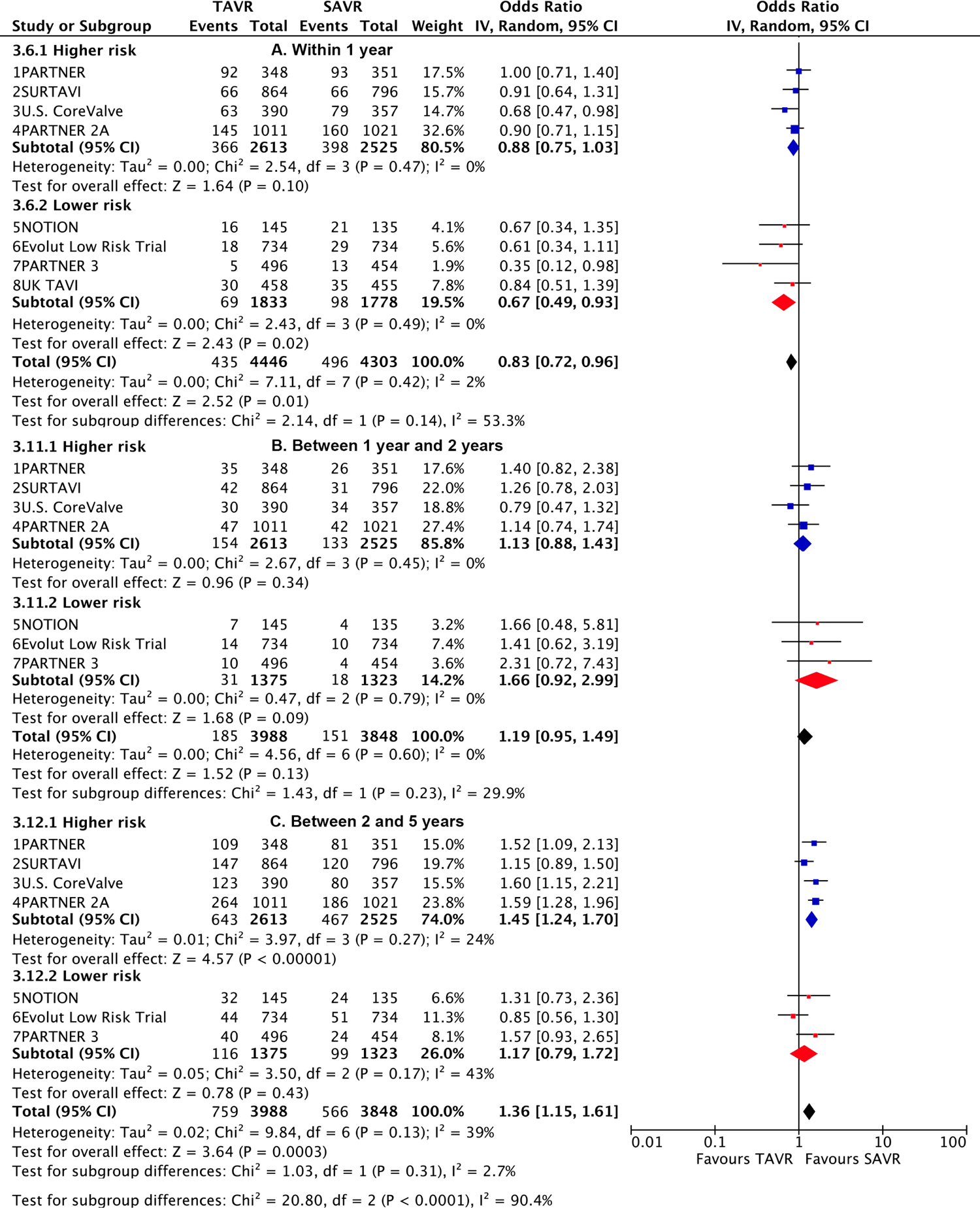
Risk estimates of all-cause death or disabling stroke for TAVR vs SAVR stratified by surgical risks according to different timing intervals. TAVR: transcatheter aortic valve replacement; SAVR: surgical aortic valve replacement (SAVR).

Subgroup analysis revealed a higher risk of long-term primary outcome in TAVR compared to SAVR among participants with higher risk (1.25 [1.07–1.47]), but no statistical difference was found in patients with lower risk (1.00 [0.77–1.29]). The higher risk of TAVR in higher-risk patients was primarily attributed to events occurring beyond 2 years (1.45 [1.24–1.7]) (p for interaction<0.0001) (Figure 2 and Table S6). The lower risk of TAVR over SAVR in lower-risk patients within 1 year (0.67 [0.49–0.93]) was not observed at long-term follow-up, and a significant temporal interaction was detected (p for interaction=0.01) (Table S7).

Subgroup analysis demonstrated a higher risk of long-term primary outcome in TAVR using BEV compared to SAVR (1.38 [1.2–1.6]), but no statistical difference was found with SEV (1.03 [0.89– 1.19]) (Figure 3). A significant interaction was observed between two valve systems (p for interaction=0.005, Table S8). The higher risk of TAVR with BEV was primarily attributed to events occurring beyond 2 years (1.57 [1.32–1.86]) (p for interaction=0.004) (Figure 4 and Table S9). The benefit of TAVR with SEV over SAVR within 1 year (0.75 [0.6–0.94]) was not observed at long-term follow-up (1.21 [0.93–1.56]), and a significant temporal interaction was detected (p for interaction=0.015) (Table S10).

**Figure 3.**
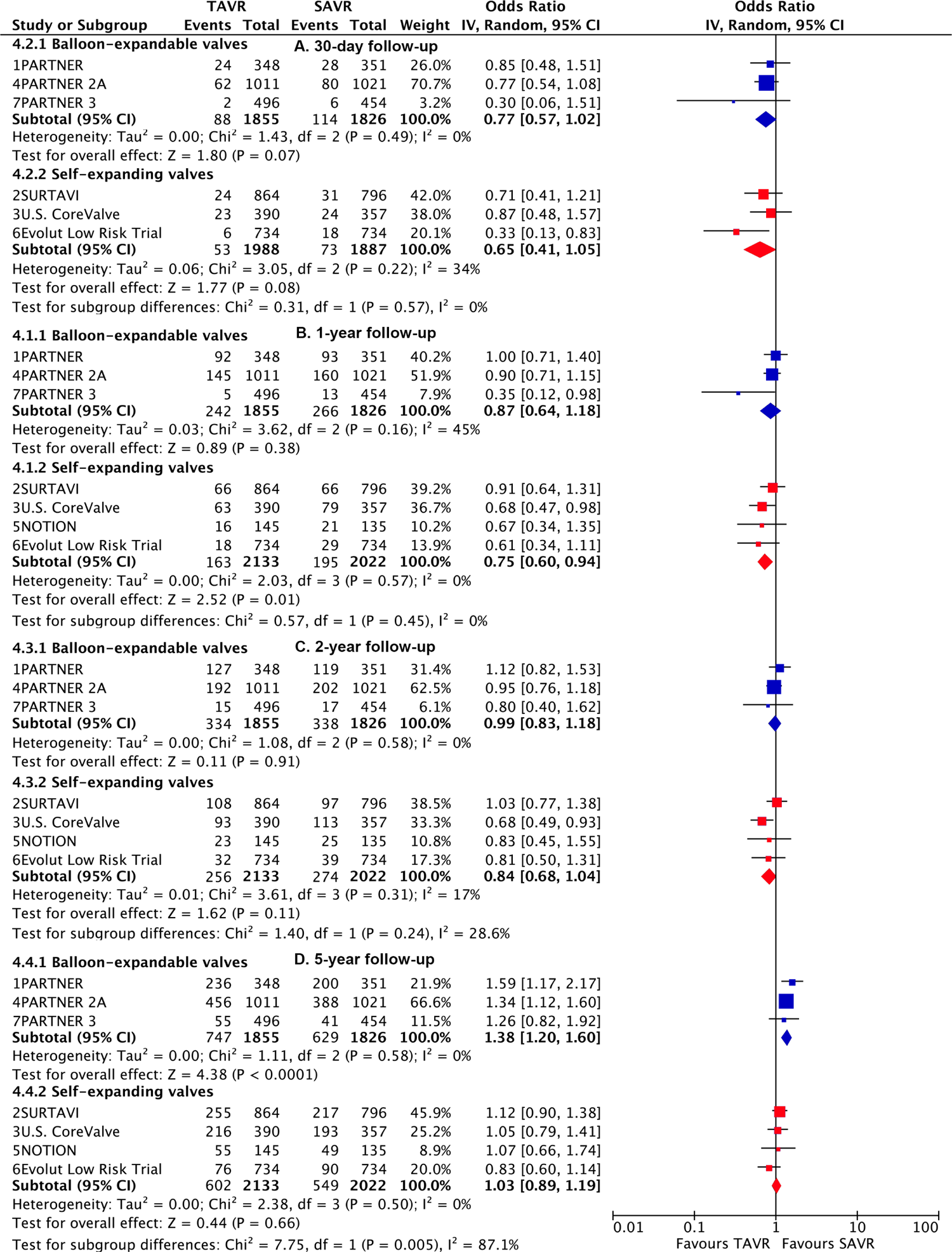
Risk estimates of all-cause death or disabling stroke for TAVR vs SAVR stratified by TAVR valve systems at different lengths of follow-up. TAVR: transcatheter aortic valve replacement; SAVR: surgical aortic valve replacement (SAVR).

**Figure 4.**
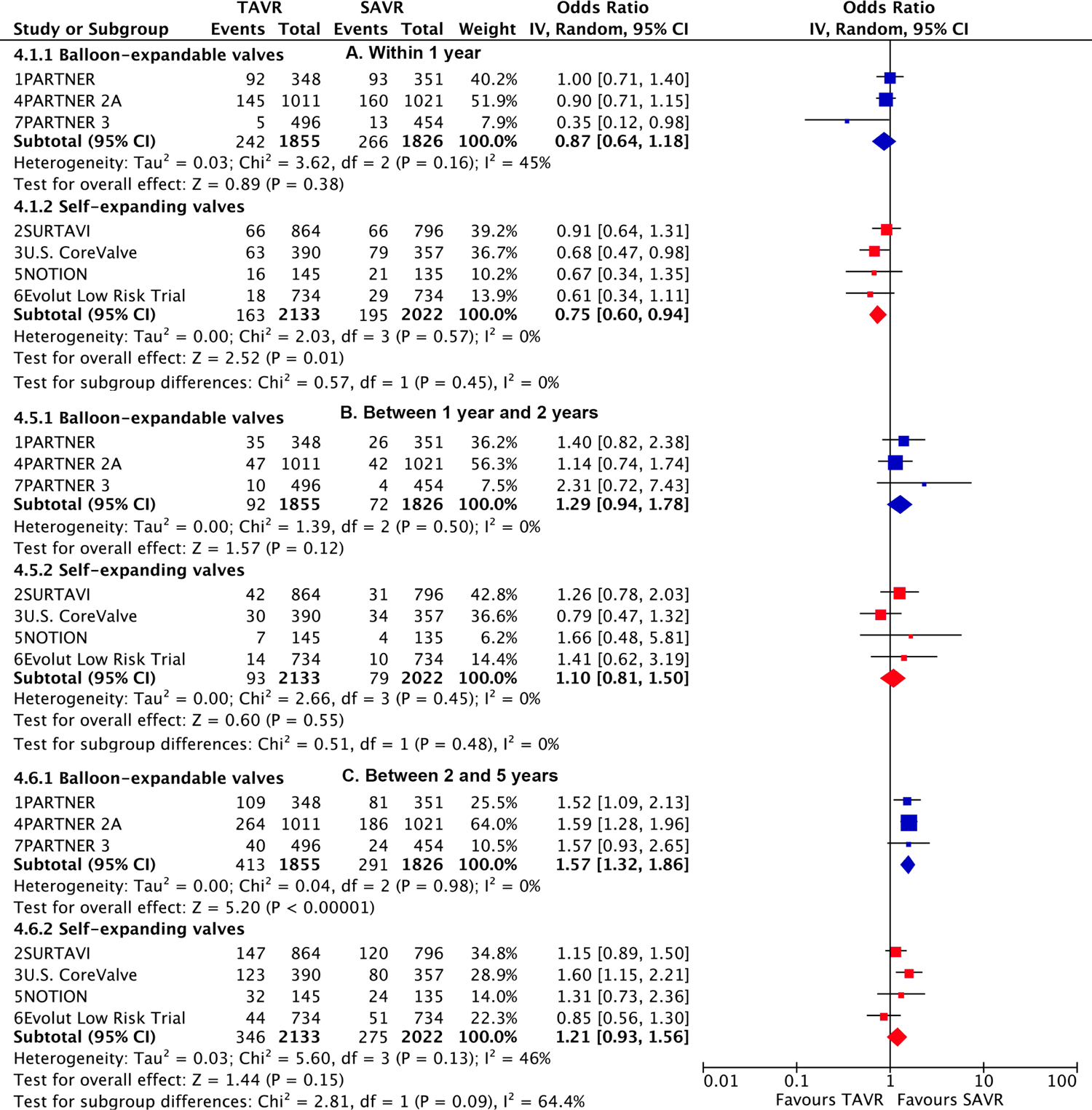
Risk estimates of all-cause death or disabling stroke for TAVR vs SAVR stratified by TAVR valve systems according to different timing intervals. TAVR: transcatheter aortic valve replacement; SAVR: surgical aortic valve replacement (SAVR).

### Other outcomes

Overall and subgroup analysis for all-cause death generated largely similar results with the primary outcome (Figure S2 and S3). At long-term follow-up, TAVR was found to have a numerically higher risk of cardiovascular death, a significantly higher risk of TIA, MVC, PPM, reintervention, rehospitalization, and moderate to severe PVL, compared to SAVR. However, TAVR showed a significantly lower risk of major bleeding and new-onset atrial fibrillation, and a comparable risk of stroke and myocardial infarction (Table 1).

**Table 1.** Other Outcomes at different durations of follow-up for TAVR compared to SAVR.

| **Outcome or Subgroup** | **Studies** | **TAVR** | **SAVR** | **OR (95% CI)** | **P value** | **I<sup>2</sup>, %** |
| --- | --- | --- | --- | --- | --- | --- |
| **All-cause death** |  |  |  |  |  |  |
| 30-day follow-up | 8 | 100/4446 | 116/4303 | 0.83 (0.61, 1.12) | 0.23 | 9 |
| 1-year follow-up | 8 | 366/4446 | 401/4303 | 0.88 (0.75, 1.02) | 0.09 | 0 |
| 2-year follow-up | 7 | 514/3988 | 522/3848 | 0.95 (0.83, 1.08) | 0.43 | 0 |
| Long-term follow-up | 7 | 1268/3988 | 1101/3848 | 1.18 (1.03, 1.36) | 0.02 | 35 |
| **Cardiovascular death** |  |  |  |  |  |  |
| 30-day follow-up | 8 | 89/4446 | 93/4303 | 0.93 (0.69, 1.25) | 0.633 | 0 |
| 1-year follow-up | 8 | 233/4446 | 257/4303 | 0.87 (0.73, 1.05) | 0.151 | 0 |
| 2-year follow-up | 7 | 320/3988 | 329/3848 | 0.94 (0.79, 1.1) | 0.427 | 0 |
| Long-term follow-up | 7 | 755/3988 | 675/3848 | 1.11 (0.99, 1.26) | 0.078 | 0 |
| **Myocardial infarction** |  |  |  |  |  |  |
| 30-day follow-up | 8 | 42/4446 | 55/4303 | 0.72 (0.48, 1.09) | 0.122 | 0 |
| 1-year follow-up | 8 | 78/4446 | 83/4303 | 0.91 (0.66, 1.25) | 0.556 | 0 |
| 2-year follow-up | 7 | 94/3988 | 96/3848 | 0.96 (0.72, 1.29) | 0.805 | 0 |
| Long-term follow-up | 7 | 198/3988 | 157/3848 | 1.12 (0.79, 1.59) | 0.514 | 51 |
| **Stroke** |  |  |  |  |  |  |
| 30-day follow-up | 8 | 160/4446 | 184/4303 | 0.85 (0.62, 1.16) | 0.301 | 41 |
| 1-year follow-up | 8 | 240/4446 | 246/4303 | 0.96 (0.7, 1.3) | 0.777 | 55 |
| 2-year follow-up | 7 | 265/3988 | 280/3848 | 0.9 (0.7, 1.15) | 0.407 | 41 |
| Long-term follow-up | 6 | 339/3254 | 325/3114 | 0.99 (0.84, 1.18) | 0.953 | 8 |
| **Transient ischemic attack** |  |  |  |  |  |  |
| 30-day follow-up | 7 | 34/3988 | 23/3848 | 1.45 (0.83, 2.52) | 0.190 | 0 |
| 1-year follow-up | 7 | 84/3988 | 60/3848 | 1.35 (0.97, 1.89) | 0.078 | 0 |
| 2-year follow-up | 6 | 99/3254 | 64/3114 | 1.49 (1.08, 2.06) | 0.014 | 0 |
| Long-term follow-up | 5 | 128/2758 | 94/2660 | 1.32 (1, 1.73) | 0.046 | 0 |
| **Major bleeding** |  |  |  |  |  |  |
| 30-day follow-up | 8 | 427/4446 | 980/4303 | 0.35 (0.18, 0.69) | 0.003 | 96 |
| 1-year follow-up | 6 | 408/3437 | 944/3372 | 0.36 (0.23, 0.56) | <0.0001 | 90 |
| 2-year follow-up | 4 | 384/2483 | 769/2463 | 0.46 (0.25, 0.84) | 0.012 | 93 |
| Long-term follow-up | 2 | 207/738 | 247/708 | 0.71 (0.57, 0.89) | 0.003 | 0.3 |
| **Major vascular complications** |  |  |  |  |  |  |
| 30-day follow-up | 8 | 286/4446 | 118/4303 | 2.74 (1.74, 4.31) | <0.0001 | 69 |
| 1-year follow-up | 6 | 236/3437 | 115/3372 | 2.31 (1.48, 3.6) | <0.0001 | 67 |
| 2-year follow-up | 4 | 181/2483 | 101/2463 | 2.03 (1.2, 3.41) | 0.008 | 71 |
| Long-term follow-up | 2 | 68/738 | 21/708 | 3.39 (2.05, 5.6) | <0.0001 | 0 |
| **Permanent pacemaker implantation** |  |  |  |  |  |  |
| 30-day follow-up | 8 | 652/4446 | 248/4303 | 2.66 (1.64, 4.31) | <0.0001 | 88 |
| 1-year follow-up | 7 | 495/3582 | 244/3507 | 2.29 (1.42, 3.7) | 0.001 | 86 |
| 2-year follow-up | 7 | 746/3988 | 319/3848 | 2.57 (1.54, 4.27) | <0.0001 | 91 |
| Long-term follow-up | 7 | 852/3988 | 395/3848 | 2.37 (1.53, 3.68) | <0.0001 | 90 |
| **New-onset atrial fibrillation** |  |  |  |  |  |  |
| 30-day follow-up | 7 | 381/3988 | 1236/3848 | 0.22 (0.16, 0.3) | <0.0001 | 83 |
| 1-year follow-up | 6 | 332/3124 | 936/3052 | 0.27 (0.18, 0.41) | <0.0001 | 88 |
| 2-year follow-up | 4 | 246/2042 | 627/1967 | 0.26 (0.16, 0.41) | <0.0001 | 86 |
| Long-term follow-up | 4 | 330/2386 | 804/2344 | 0.28 (0.2, 0.38) | <0.0001 | 75 |
| **Moderate to severe paravalvular leak** |  |  |  |  |  |  |
| 30-day follow-up | 7 | 166/3438 | 14/3465 | 11.41 (6.69, 19.47) | <0.0001 | 0 |
| 1-year follow-up | 7 | 115/3040 | 15/2494 | 5.67 (3.25, 9.88) | <0.0001 | 0 |
| 2-year follow-up | 6 | 127/1976 | 16/1707 | 7.97 (2.21, 28.77) | 0.002 | 69 |
| Long-term follow-up | 6 | 49/1695 | 3/1449 | 7.94 (3.12, 20.22) | <0.0001 | 0 |
| **Reintervention** |  |  |  |  |  |  |
| 30-day follow-up | 5 | 22/4098 | 6/3952 | 2.85 (1.16, 7) | 0.022 | 0 |
| 1-year follow-up | 6 | 54/4098 | 19/3952 | 2.48 (1.45, 4.23) | 0.001 | 0 |
| 2-year follow-up | 4 | 47/2906 | 14/2763 | 2.92 (1.3, 6.55) | 0.009 | 35 |
| Long-term follow-up | 6 | 82/3640 | 42/3497 | 1.86 (1.05, 3.28) | 0.032 | 46 |
| **Rehospitalization** |  |  |  |  |  |  |
| 30-day follow-up | 5 | 130/3453 | 153/3356 | 0.78 (0.56, 1.1) | 0.157 | 40 |
| 1-year follow-up | 6 | 393/3843 | 378/3713 | 0.97 (0.74, 1.27) | 0.828 | 65 |
| 2-year follow-up | 6 | 528/3843 | 450/3713 | 1.12 (0.88, 1.43) | 0.371 | 66 |
| Long-term follow-up | 6 | 825/3843 | 658/3713 | 1.23 (1.0, 1.5) | 0.047 | 65 |

TAVR: transcatheter aortic valve replacement; SAVR: surgical aortic valve replacement (SAVR).

The increased risk of TAVR on cardiovascular death was primarily attributed to events occurring beyond 2 years, rehospitalization attributed to events beyond 1 year, while TIA, MVC, and reintervention were primarily associated with events within 1 year. The benefits of TAVR on major bleeding and new-onset atrial fibrillation were mainly attributed to lower events occurring within 1 year. The risk of PPM at long-term follow-up was primarily attributed to higher events occurring within 1 year in TAVR, with the risk attenuating but still higher in TAVR between 1 year and 2 years and beyond 2 years (Table S5).

In subgroup analysis, a statistically higher risk of long-term reintervention and rehospitalization was observed in TAVR compared to SAVR among participants at higher risk, while no statistical difference was found in patients at lower risk (Table S11). Significant interaction was detected between the two risk groups (both p for interaction <0.0001). The lower risk of rehospitalization in TAVR over SAVR in lower-risk patients within the first year was not observed during long-term follow-up (Table S7). Subgroup analysis indicated a statistically higher risk of long-term PPM in TAVR compared to SAVR, regardless of participants’ higher or lower risk. The SEV showed a higher risk than the BEV, with a significant difference (p for interaction<0.0001) (Table S8). Pooled analyses of all outcomes stratified by surgical risks and TAVR systems at 30-day, 1-year, and 2-year follow-up, are presented in Table S12-S17.

### Sensitivity analyses

The analysis of primary outcome using the Hartung-Knapp-Sidik-Jonkman variance correction and excluding each trial one time revealed largely similar findings (Figure S4-S7).

## Discussion

This present meta-analysis, including comprehensive data from all available trials comparing TAVR with SAVR, with >8,000 patients and long-term follow-up data from nearly all trials, yields several important conclusions. First, TAVR was associated with a higher risk of long-term primary outcome compared to SAVR among participants with higher risk, but not among those with lower risk. However, a significant temporal interaction was detected in both risk profiles. Second, TAVR with BEV was associated with a higher risk of long-term primary outcome compared to SAVR, whereas no statistical difference was found with SEV. There was a significant interaction between the two valve systems, and a temporal interaction was observed in both TAVR systems. Third, landmark analysis revealed a lower risk of primary outcome in TAVR compared to SAVR within the initial 30 days, comparable between 30 days to 2 years, and a significant higher risk beyond 2 years. Fourth, overall analysis showed that TAVR was associated with a higher long-term risk of all-cause death, TIA, MVC, PPM, reintervention, rehospitalization, and moderate to severe PVL, a comparable risk of stroke and myocardial infarction, but a lower risk of major bleeding and new-onset atrial fibrillation.

We conducted a comprehensive search on PubMed to identify relevant meta-analyses comparing the long-term outcomes of TAVR and SAVR. However, these meta-analyses included 3 to 4 trials with 5-year follow-up data, focusing exclusively on patients with higher risks [2, 24, 25]. In contrast, our meta-analysis incorporated a larger dataset, comprising 7 trials with long-term follow-up data, encompassing both higher- and lower-risk patients. It is important to note that our study utilized long-term data from nearly all registered large RCTs. One of the identified meta-analyses employed a network meta-analysis approach but considered 1-to-2-year follow-up as long-term [26]. Another meta-analysis included only 3 RCTs but supplemented them with 7 propensity-score matching observational studies, which were limited by inadequate adjustment for important confounding [27]. We also performed several additional analyses. Firstly, we conducted a landmark analysis to assess the differences in TAVR outcomes within specific time intervals, revealing significant temporal variations in the effect of TAVR. Secondly, we conducted subgroup analyses based on TAVR systems and surgical risks, revealing noteworthy distinctions between subgroups.

None of the trials included were specifically designed to have sufficient statistical power to detect a significant reduction in all-cause death. However, our meta-analysis revealed a significant higher risk of long-term mortality associated with TAVR. This finding aligns with the temporal trend observed in primary outcome. Further subgroup analysis indicated a significantly higher risk of all-cause death in TAVR among higher-risk patients and with BEV, but no significant difference was observed in lower-risk patients or with SEV. Importantly, the temporal trend was also only evident in the former two subgroups. A separate meta-analysis of 7 propensity matched studies corroborated our findings by showing a significantly higher risk of mortality at 5-year follow-up [27].

TAVR demonstrated initial superiority over SAVR within the first year but lost this advantage thereafter in lower risk patients. Given that lower risk patients typically have good life expectancy, this temporal interaction warrants intensive and close attention. In the PARTNER 3 trial, Kaplan-Meier event curves for the primary outcome crossed around the 2- to 3-year mark, thereafter favoring SAVR, while in the Evolut Low Risk trial, the curves remained parallel, favoring TAVR [8, 9]. Although there were some differences, the pooled analysis of long-term data from these lower-risk trials did not show substantial heterogeneity (*I^2^*=20%). A large real-world registry including 42,586 patients who underwent isolated SAVR and meeting the inclusion and exclusion criteria for the PARTNER 3 and Evolut Low Risk trials, revealed excellent survival rates in low-risk patients following SAVR, with all-cause mortality of 7.1% at 5 years and 12.4% at 8 years [28]. Similar findings were observed in other large registries [29]. Determining whether TAVR can achieve such excellent long-term outcomes as SAVR will require robust evidence from follow-up periods exceeding 10 years. The recommendation of TAVR in these patients is pending this evidence.

We showed a higher long-term risk of primary outcome and all-cause death in TAVR compared to SAVR among higher-risk patients. These observations seem a paradox, *i.e.* patients with a higher surgical risk actually had better long-term outcomes when they underwent surgery instead of opting for TAVR. Notably, the short-term risk of all-cause death was not decreased in TAVR in higher risk patients. This observation was similar to several meta-analyses with higher-risk patients [2, 24].

However, these trials utilized early-generation TAVR valves and were performed in earlier years, potentially involving less mature implantation technologies and suboptimal antithrombotic medications. Unfortunately, no randomized trials in high-risk patients using newer-generation valves have been conducted thus far. There have been some propensity-matched studies that shed light on this topic. For instance, a study involving 72 pairs of high-risk patients, although utilizing mixed generations of TAVR valves, showed a lower in-hospital mortality rate but a higher risk of all-cause death at 5-year follow-up in the TAVR group [30]. Another propensity-matched analysis of 783 pairs of intermediate-risk patients (mean age: 81.7 years, mean STS score: 5.5) using newer-generation SAPIEN 3 valves demonstrated a comparable risk of death or disabling stroke at 5 years compared to SAVR [31]. It’s worth noting that this intermediate-risk category falls into the higher-risk group as per our study’s classification. Further studies are warranted to evaluate the performance of TAVR with newer-generation valves compared to SAVR in the context of higher-risk patients.

An interesting finding of our analysis was the significant interaction between BEV and SEV regarding the primary outcome and all-cause death at long-term follow-up. A temporal interaction was observed in BEV for both the primary outcome and all-cause death, while in SEV, it was observed only for the primary outcome. These temporal trends closely align with those reported in the PARTNER 2A trial [5], which compared early-generation BEV TAVR with SAVR in higher surgical risk patients, and the PARTNER 3 trial [9], which compared newer-generation BEV TAVR with SAVR in lower surgical risk patients. Landmark analyses of clinical events between 2 and 5 years in both trials demonstrated higher rates of all-cause death and the primary outcome in TAVR compared to SAVR. Similarly, in another trial of BEV TAVR, the Kaplan-Meier event curves for all-cause death converged at 2 years [4]. In contrast, trials comparing SEV TAVR to SAVR showed Kaplan-Meier event curves for the primary endpoint that remained parallel, favoring TAVR in the Evolut Low Risk trial [8], nearly overlapped in the SURTAVI [6] and NOTION [20] trials, and converged until the 5-year mark in the U.S. CoreValve trial [3]. Long-term data from head-to-head comparisons of BEV with SEV TAVR have been reported in only one RCT [32]. In this trial, with 241 high-risk patients randomly assigned to early generation BEV and SEV, all-cause mortality (53.4% vs. 47.6%) and cardiovascular mortality (31.6% vs. 21.5%) at 5 years were numerically higher in the BEV group compared with the SEV group, consistent with our findings. These differences might be attributed to better forward flow hemodynamics and less structural valve deterioration in SEV compared to BEV [32]. Several propensity-matched studies showed varied findings, but these conclusions were limited by residual confounders that could not be fully accounted for, such as patients’ anatomical suitability. It is likely that more patients with extensive outflow tract calcifications, low implanted coronary arteries, or complex and small femoral access received SEV [33]. We found no significant difference between BEV and SEV at short-term follow-up, which is also consistent with findings from other RCTs [34, 35].

Our analysis had several strengths. Firstly, we incorporated the largest number of RCTs with long-term follow-up outcomes, ensuring a comprehensive evaluation of the data. Additionally, the trials included in our analysis had nearly identical follow-up durations, enabling landmark analyses and mitigating the potential impact of variations in follow-up durations on the outcomes. Furthermore, there was minimal heterogeneity observed across trials for both the primary outcomes and all death outcomes across all follow-up durations.

However, it is important to acknowledge some limitations. Firstly, our analysis was based on trial-level rather than patient-level data. Although we performed subgroup analyses based on clinically relevant subgroups, we were unable to conduct more detailed meta-regression analyses to account for potential confounding factors beyond the subgroup variables. Secondly, the trials involving high-risk patients primarily utilized early-generation TAVR valves and were conducted in earlier years, therefore, we were unable to fully account for the potential impact of learning curves and advancements in TAVR valve technology on the observed outcomes. Thirdly, concomitant procedures were performed in both TAVR and SAVR groups in original trials, which could potentially influence the evaluation of isolated TAVR versus isolated SAVR. Fourthly, our assessment of publication bias was limited by the relatively small number of trials, potentially affecting the ability to detect small-study effects.

## Conclusions

TAVR was associated with a higher long-term risk of primary outcome compared to SAVR in higher-risk patients and with balloon-expandable valves. However, a characteristic temporal interaction was documented in all subgroups. Longer-term follow-up data from low-risk trials and large trials comparing TAVR with balloon-expandable and self-expanding valves are warranted to test these findings.

## Abbreviations

MVC: major bleeding, major vascular complication

OR: odds ratio

RCT: randomized controlled trials

SAVR: surgical aortic valve replacement

TAVR: Transcatheter aortic valve replacement

TIA: transient ischemic attack

PPM: permanent pacemaker implantation

PVL: paravalvular leak.

## Competing interests

The authors declare that they have no competing interests.

## Funding

None.

## Authors’ contributions

XWZ and LK selected studies and extracted the data, analyzed and interpreted the data, wrote the first draft of the manuscript. LW, KW, and WX contributed to the study protocol and interpreted the data. XLZ and BX conceived the study, interpreted the data, and wrote the first draft of the manuscript. All authors read and approved the final manuscript.

## Data Availability

Data are available upon reasonable request from Dr. Xinlin Zhang.

## Acknowledgements

None.

## Notes

### Competing Interest Statement

The authors have declared no competing interest.

